# Neutrophil-to-Lymphocyte Ratio Predicts Severe Illness Patients with 2019 Novel Coronavirus in the Early Stage

**DOI:** 10.1101/2020.02.10.20021584

**Authors:** Jingyuan Liu, Yao Liu, Pan Xiang, Lin Pu, Haofeng Xiong, Chuansheng Li, Ming Zhang, Jianbo Tan, Yanli Xu, Rui Song, Meihua Song, Lin Wang, Wei Zhang, Bing Han, Li Yang, Xiaojing Wang, Guiqin Zhou, Ting Zhang, Ben Li, Yanbin Wang, Zhihai Chen, Xianbo Wang

## Abstract

**Background:** Severe ill patients with 2019 novel coronavirus (2019-nCoV) infection progressed rapidly to acute respiratory failure. We aimed to select the most useful prognostic factor for severe illness incidence.

**Methods:** The study prospectively included 61 patients with 2019-nCoV infection treated at Beijing Ditan Hospital from January 13, 2020 to January 31, 2020. Prognostic factor of severe illness was selected by the LASSO COX regression analyses, to predict the severe illness probability of 2019-CoV pneumonia. The predictive accuracy was evaluated by concordance index, calibration curve, decision curve and clinical impact curve.

**Results:** The neutrophil-to-lymphocyte ratio (NLR) was identified as the independent risk factor for severe illness in patients with 2019-nCoV infection. The NLR had a c-index of 0.807 (95% confidence interval, 0.676–0.38), the calibration curves fitted well, and the decision curve and clinical impact curve showed that the NLR had superior standardized net benefit. In addition, the incidence of severe illness was 9.1% in age ≥ 50 and NLR < 3.13 patients, and half of patients with age ≥ 50 and NLR ≥ 3.13 would develop severe illness. Based on the risk stratification of NLR with age, the study developed a 2019-nCoV pneumonia management process.

**Conclusions:** The NLR was the early identification of risk factors for 2019-nCoV severe illness. Patients with age ≥ 50 and NLR ≥ 3.13 facilitated severe illness, and they should rapidly access to intensive care unit if necessary.

## Introduction

Coronavirus was a large virus family, which was known to cause common cold and serious illness such as Middle East respiratory syndrome (MERS) and severe acute respiratory syndrome (SARS).^1-4^ It was found that the 2019 novel coronavirus (2019-nCoV) was the cause of unexplained viral pneumonia in Wuhan, China in December 2019, and was recognized by the World Health Organization (WHO) on January 12, 2020. In the following month, 2019-nCoV was transmitted in Hubei Province, throughout China and even other countries,^5^ causing 34,662 confirmed cases by February 8, 2020.

Most patients with novel coronavirus infection were mild. Moderate illness often experienced dyspnea after one week. Severe ill patients progressed rapidly to acute respiratory failure, acute respiratory distress syndrome, metabolic acidosis, coagulopathy, and septic shock. Early identification of risk factors for severe illness facilitated appropriate supportive care and promptly access to the intensive care unit (ICU) if necessary. For mild patients, general isolation and symptomatic treatment were available, and ICU-care was needed unless the condition worsens rapidly, such to reduce the mortality and alleviate the shortage of medical resources.

We aimed to compare the clinical characteristics, imaging features, treatments, and outcomes of patients with mild and moderate or severe 2019-nCoV infection, to explore the most useful prognostic factor for accurate individualized assessment of the severe incidence of 2019-nCoV infection.

## Methods

### Patient selection

This study was a prospective single-center study, which included 61 patients with 2019-nCoV infection treated at Beijing Ditan Hospital from January 13, 2020 to January 31, 2020. Diagnosis of 2019-nCoV pneumonia and clinical classification according to the new coronavirus pneumonia diagnosis and treatment plan (trial version 4) developed by the National Health Committee of the People’s Republic of China (http://www.nhc.gov.cn/). The clinical classifications are as follows: (1) mild, with fever, respiratory tract symptoms, and imaging shows pneumonia. (2) moderate, meet any of the following: a) respiratory distress, respiratory rate ≥ 30 beats / min; b) in the resting state, means oxygen saturation ≤ 93%; c) arterial blood oxygen partial pressure / oxygen concentration ≤ 300mmHg (1mmHg = 0.133kPa). (3) severe, one of the following conditions: a) respiratory failure occurs and requires mechanical ventilation; b) Shock occurs; c) ICU admission is required for combined organ failure. This study was approved by the Ethics Committee of Beijing Ditan Hospital, and all patients signed the informed consent.

### Baseline data collection

All suspected 2019-nCoV infection patients were taken upper respiratory throat swab samples at admission and stored in virus transport medium, which were transported to Beijing CDC for laboratory diagnosis. Influenza A virus (H1N1, H3N2, H7N9), influenza B virus, bacterial and fungal detections of sputum or respiratory secretions were performed at this hospital. Epidemiological history, comorbidity, vital signs, symptoms and signs were recorded in detail, and laboratory tests including biochemical Indicators, blood routine, C-reactive protein, chest radiograph or CT scan.

### Follow-up

After admission, the patients were re-examined for laboratory indexes and imaging examination, and recorded symptoms and signs, treatments and outcome events. The endpoint of this study was the severe illness developed.

### Statistical analysis

Age and days were represented by median (range), categorical variables by number (%), and laboratory data by mean (interquartile range). The cutoff value of neutrophil-to-lymphocyte ratio (NLR) was calculated based on the maximum Youden index. Compared the differences between the two groups with t-test, chi-square test or Mann-Whitney U test. LASSO COX regression was used to select independent risk factors that affect outcomes. Analyses were performed using SPSS 22.0 statistical package (SPSS, Inc., Chicago, IL, USA) and R version 3.0.2 was used to establish nomogram, calibration, decision curve and clinical impact curve. P-value < 0.05 was considered statistically significant.

## Results

Of the 61 patients with 2019-nCoV infection included in this study, 44 were diagnosed as mild (mild group) and 17 as moderate or severe (severe group) on admission. The median age of the two groups was statistically different, the mild group was 41 years old and the severe group was 56 years old. None of the 61 patients had a history of Huanan seafood market exposure in Wuhan, 17 of them (27.9%) had not left Beijing recently, but had a close exposure history with 2019-nCoV pneumonia patients, and the other 44 (72.1%) patients were Wuhan citizens or visited Wuhan recently. There was no significant difference of diabetes, cardiovascular disease and chronic obstructive pulmonary disease between the two groups, but the patients with hypertension in the severe group were more than those in the mild group (p = 0.056) (Table 1). Among 61 cases, 60 (98.4%) cases had fever, 5 (8.2%) cases had high fever (> 39 °C), 3 (4.9%) cases had dyspnea. 7 (11.5%) cases had mild shortness of breath. Most of the patients in the severe group had cough, sputum production, and fatigue. 11 (18.0%) cases had gastrointestinal symptoms. Blood neutrophil count in the severe group was significantly higher than that in the mild group, blood lymphocyte count, serum sodium and chlorine levels were significantly lower, and the rate of bacterial infection was significantly increased (Table 1).

**Table 1.**
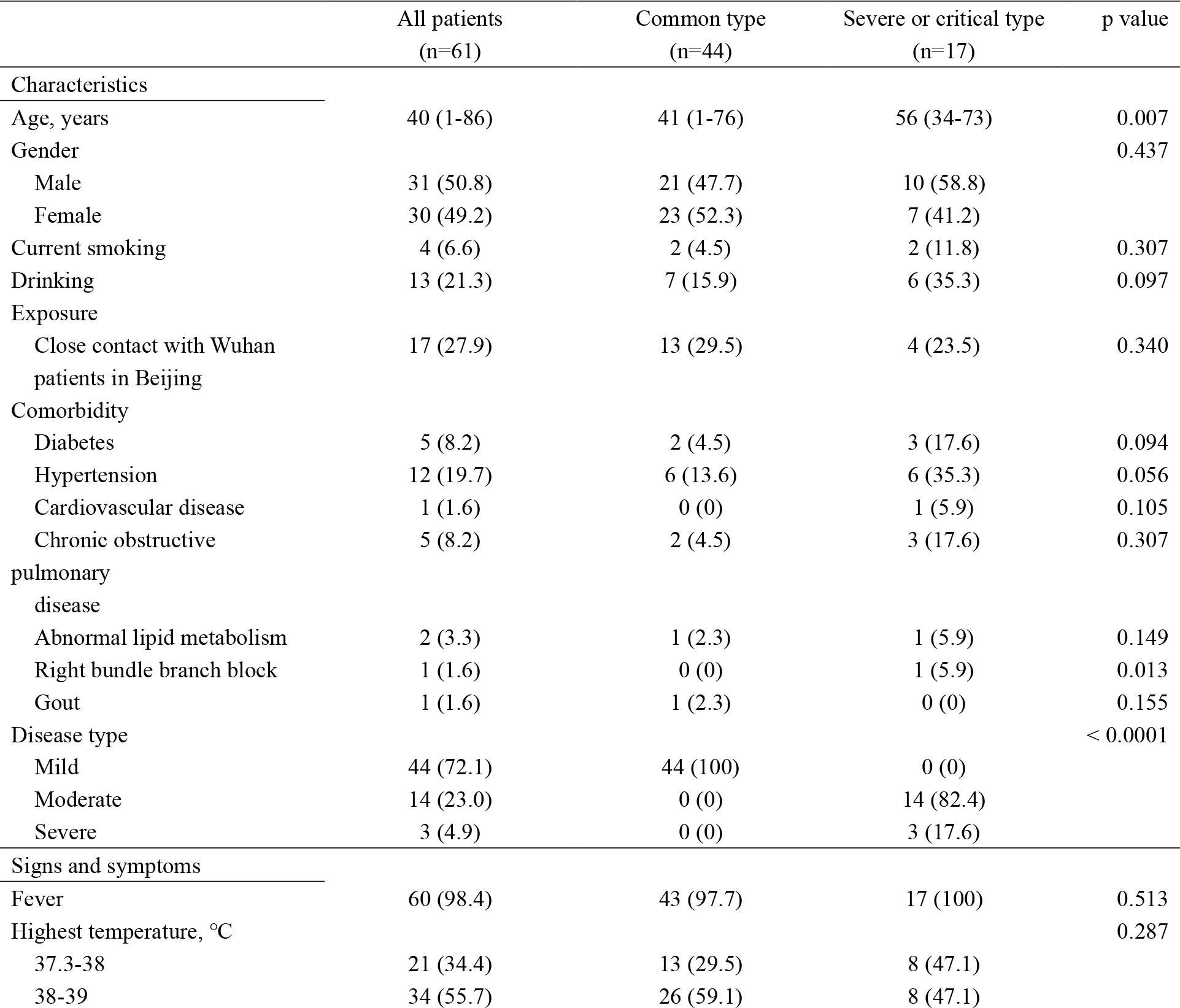
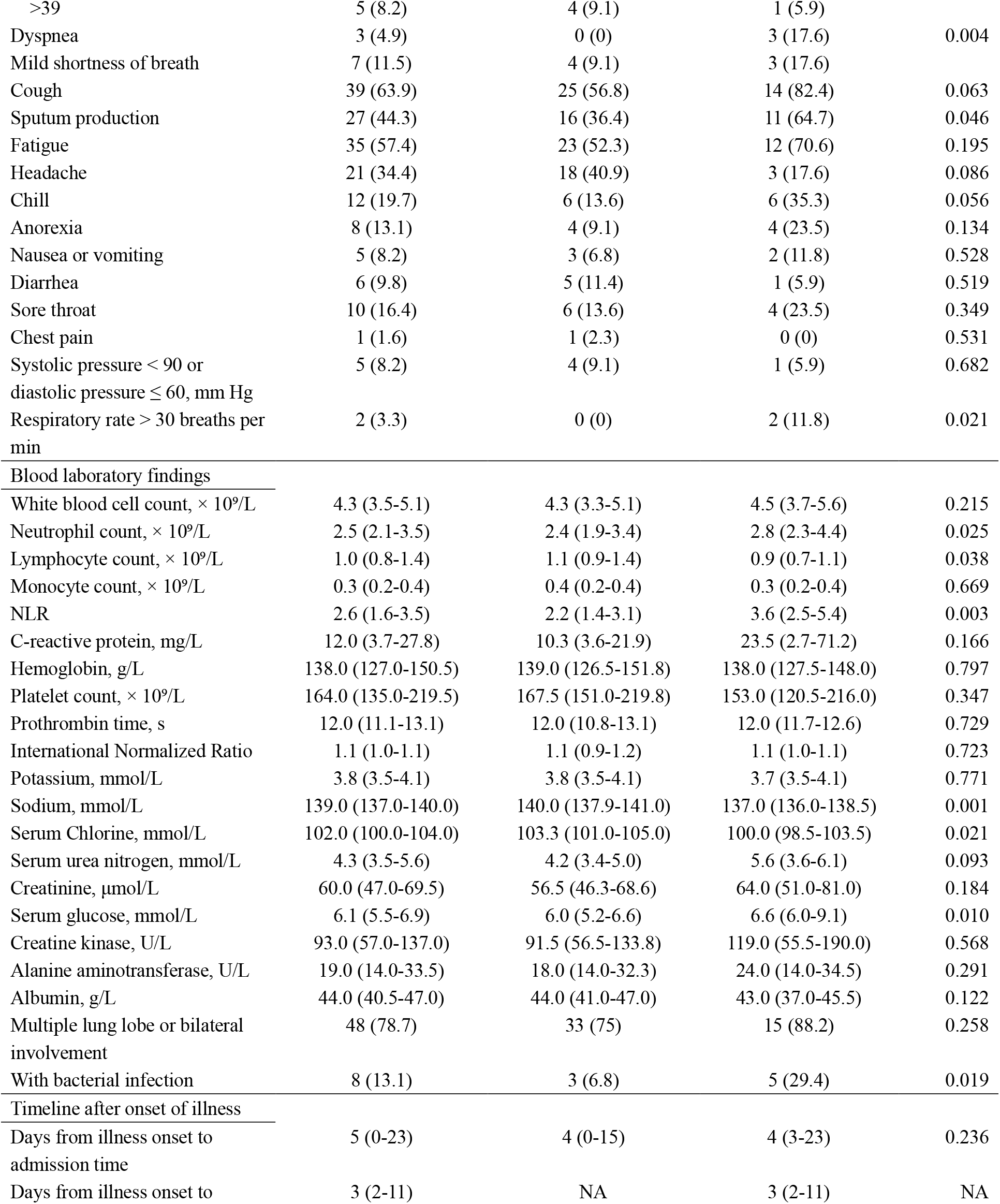
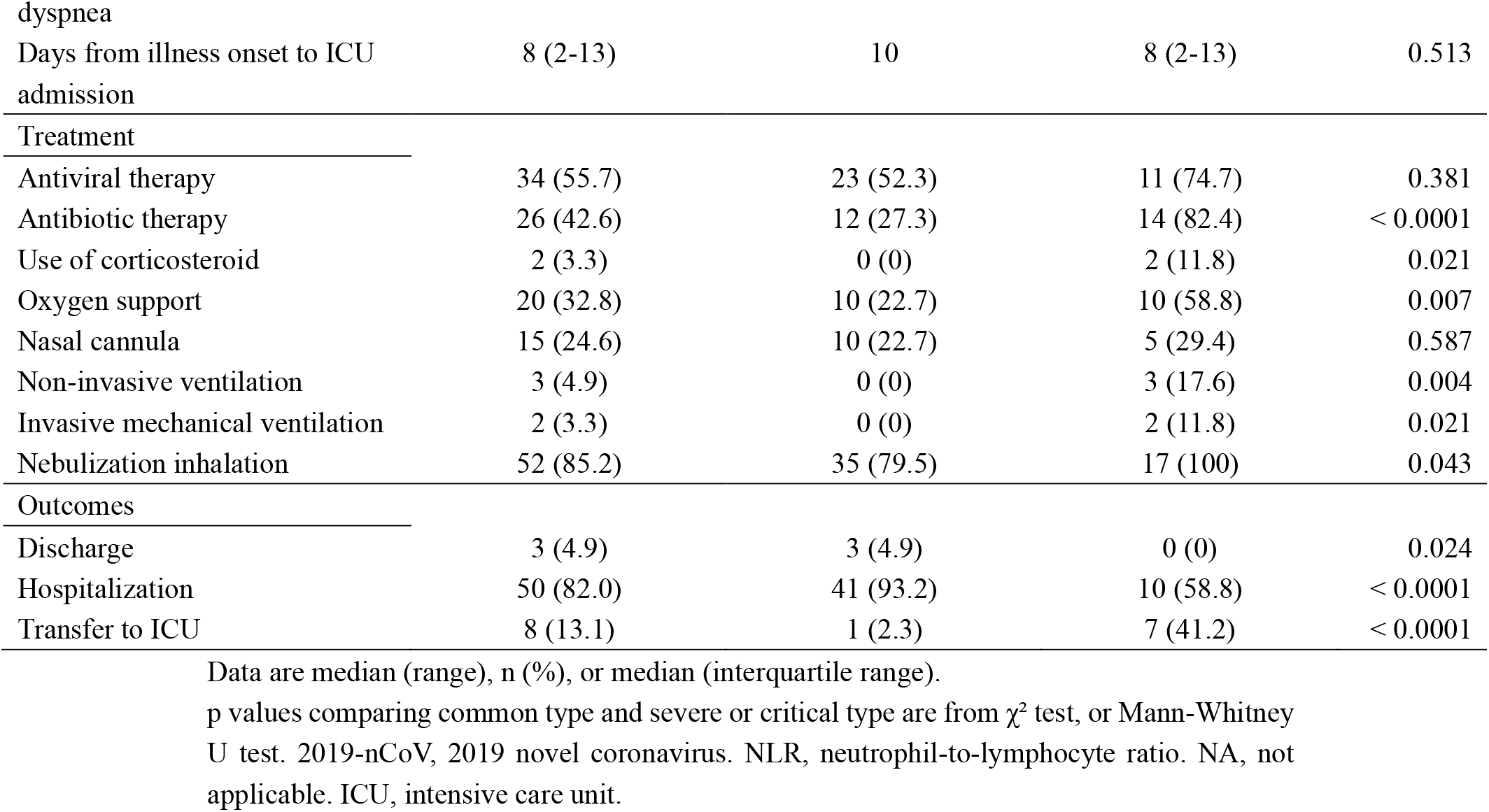
Demographics and characteristics of patients infected with 2019-nCoV

The median days from illness onset to admission were 4 days in both groups. All patients were isolated after admission, 34 (55.7%) patients received antiviral treatment, of which 8 patients received oseltamivir (75 mg every 12 h, orally), 26 patients received lopinavir and ritonavir tablets (200 mg twice daily, orally). Nearly half of the patients received antibiotic therapy (42.6%), and 84.2% of the patients in the severe group received antibiotic therapy. One patient was received methylprednisolone for 3 days before admission and stopped after admission, and another patient took methylprednisolone 8 mg every other day for ten months for optic neuromyelitis and continued taking it after admission. Most of the patients in the severe group received oxygen support and nebulization inhalation therapy, of which 3 patients received non-invasive ventilation and 2 received invasive mechanical ventilation. Nebulization inhalation drugs included recombinant human interferon α2b and acetylcysteine. By the end of Jan 31, no patients died, 3 patients were discharged, and the remaining patients were in hospital, of which 8 patients progressed to severe illness and still in the ICU (Table 1).

X-ray or CT showed multiple lung lobe or bilateral involvement in 48 (78.7%) patients. Figure 1 showed the CT images of a typical patient in early, consolidation, absorption and dissipation stages.

**Figure 1.**
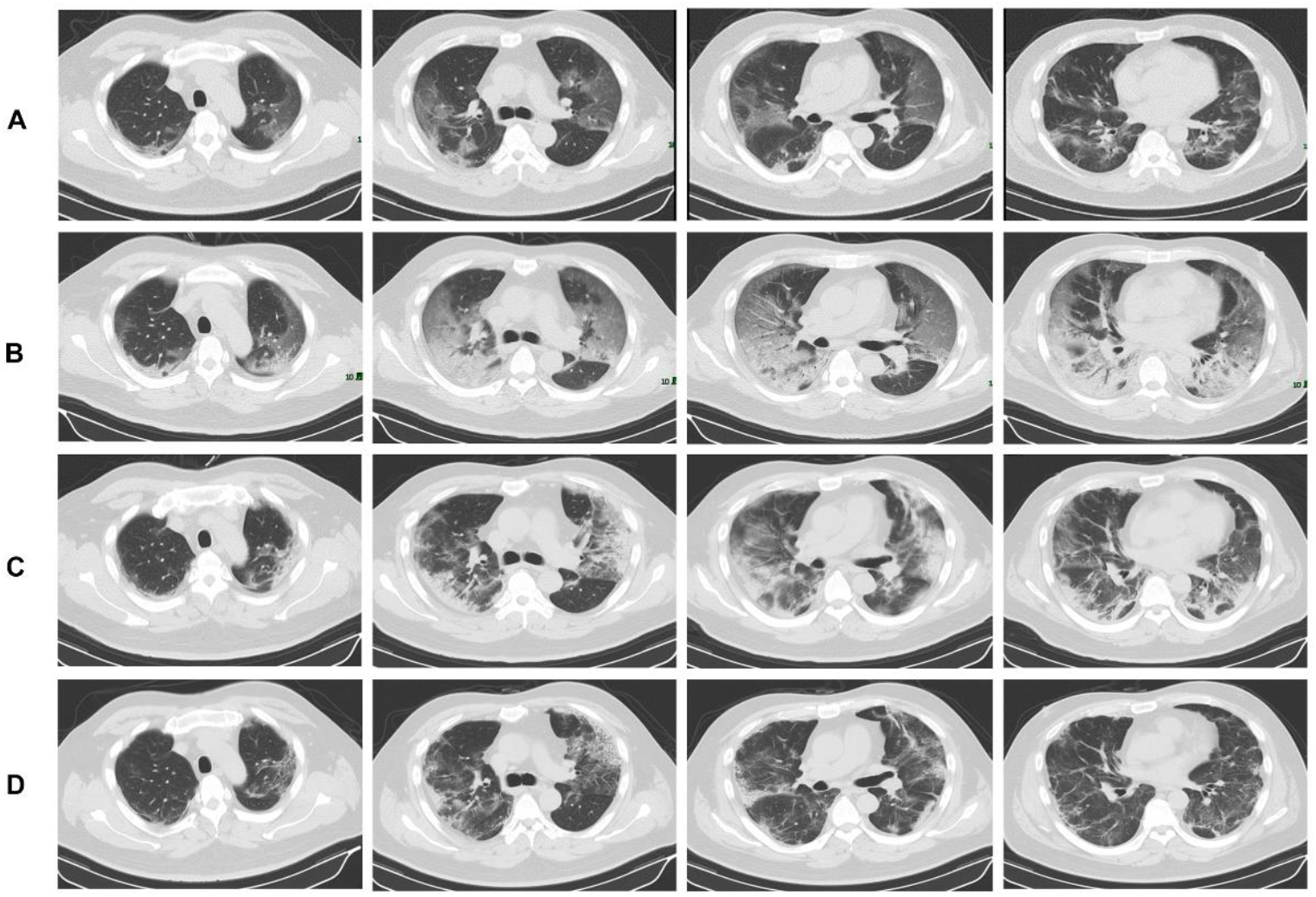
A 50-year-old man with 2019 novel coronavirus (2019-nCoV) infection. (A) Ground glass shadow in multiple lobes and segments of bilateral lungs, and the lesions were adjacent to the pleura (Illness Day 8, Hospital Day 0). (B) Ground glass shadow expanding and consolidation in bilateral lung (Illness Day 11, Hospital Day 3). (C) Ground glass shadow absorption and reduced consolidation area (Illness Day 15, Hospital Day 7). (D) Lesion dissipation (Illness Day 20, Hospital Day 12).

### Prognostic factors of severe illness

26 variables were included in the LASSO regression, the results showed that age, NLR and hypertension obtained from the 61 cases cohort were predictive factors for severe illness incidence when the partial likelihood deviance was smallest, and NLR was the significant factors when the lambda was 1 standard error (Figure 2A and 2B). The three factors mentioned above were included in the multivariate COX regression analyses, the results showed that age and NLR were predictive factors. However, the hazard ratios (HR) of age was close to 1, then age was transformed into a categorical variable (< 50 years / ≥ 50 years) based on cutoff value, and then the three variables were included in the COX regression analyses again. Finally, NLR was selected as the most useful prognostic factor affecting the prognosis for severe illness incidence by the forward selection procedure.

**Figure 2.**
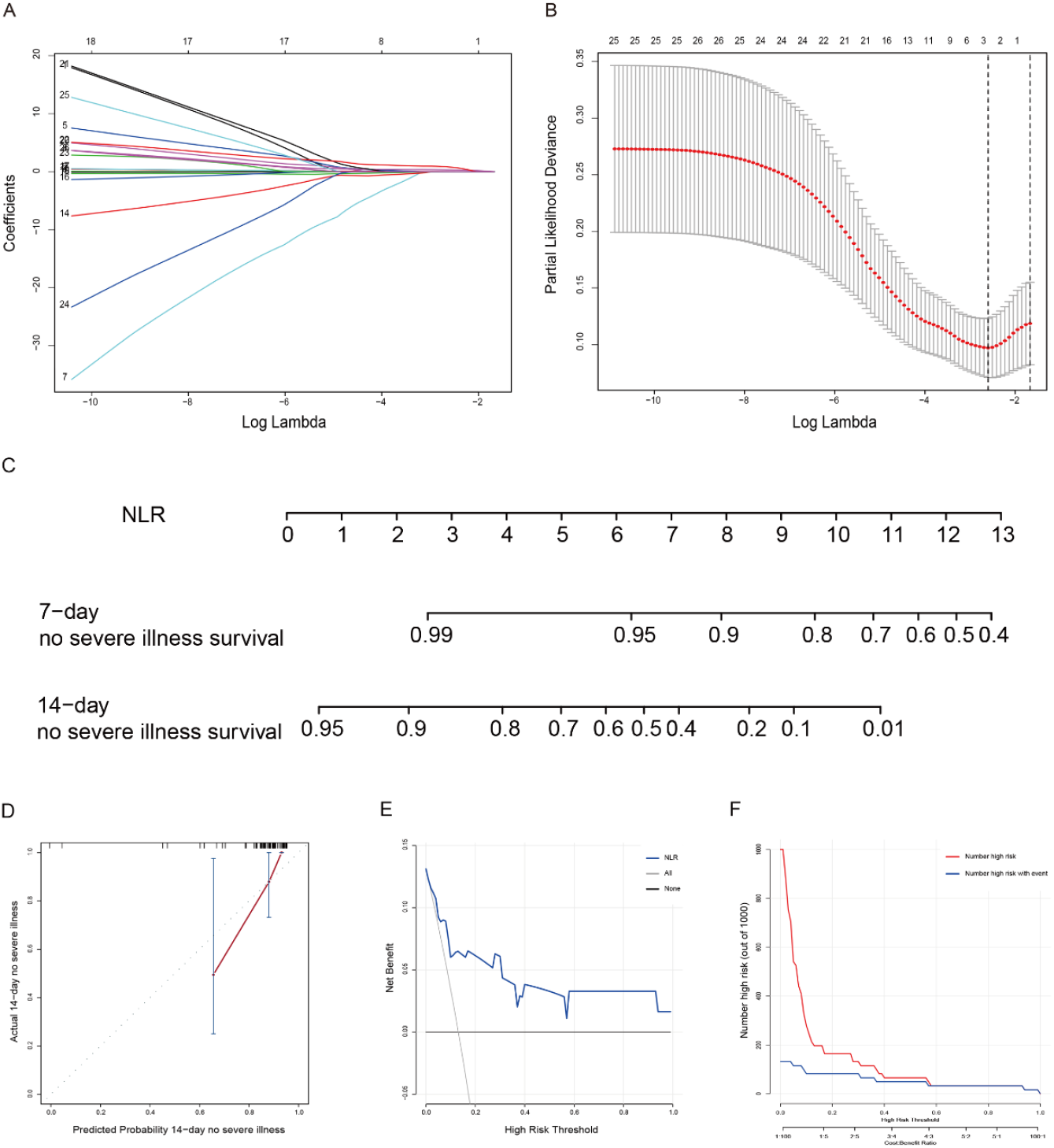
Prognostic factor neutrophil-to-lymphocyte ratio (NLR) was selected by LASSO regression analyses. (A) LASSO coefficient profiles of the non-zero variables of 2019-nCoV pneumonia. (B) Mean−Squared Error plot of the lowest point of the red curve, which corresponds to a three-variable model. The dashed line on the right was a more concise model within one standard error (the number of variables was one). (C) Nomogram predicting 7-day and 14-day no severe illness survival of patients with 2019-nCoV pneumonia. (D) Calibration plot, (E) decision curve and (F) clinical impact curve of the nomogram for no severe illness survival in the 2019-nCoV cohort, in which the predicted probability of survival was compared well with the actual survival and had superior standardized net benefit.

### Nomogram

The nomogram was established based on NLR, which were used to predict the no-severe ill survival rates of 7-day and 14-day (Figure 2C). The nomogram had a c-index of 0.807 (95% confidence interval [CI], 0.676–0.938) for predicting the severe illness probability. The calibration curves showed that the predicted rates were in agreement with the actual results observed in the cohort (Figure 2D). The decision curve and clinical impact curve showed that the NLR had superior standardized net benefit and influence on the outcome of patients (Figure 2E and 2F).

### Comparison of the NLR with MuLBSTA and CURB-65

Using receiver operating characteristic analysis, the predictive value of the NLR for the incidence of severe illness was compared to that of the MuLBSTA and CURB-6 models. The results showed that the NLR had the highest area under curve (AUC) (0.849), and had higher sensitivity and specificity than the other two models (Table 2).

**Table 2.** Predictive value of the NLR, MuLBSTA and CURB-65.

|  | AUC<br>(95% CI) | c-index<br>(95% CI) | SEN<br>(95% CI) | SPE<br>(95% CI) | PPV<br>(95% CI) | NPV<br>(95% CI) | DLR positive<br>(95% CI) | DLR negative<br>(95% CI) |
| --- | --- | --- | --- | --- | --- | --- | --- | --- |
| NLR | 0.849<br>(0.707-0.991) | 0.807<br>(0.676-0.938) | 0.875<br>(0.473-0.997) | 0.717<br>(0.577-0.832) | 0.318<br>(0.200-0.955) | 0.974<br>(0.823-0.987) | 3.092<br>(1.871-5.109) | 0.174<br>(0.028-1.100) |
| MuLBSTA | 0.762<br>(0.585-0.938) | 0.771<br>(0.659-0.883) | 0.875<br>(0.473-0.997) | 0.679<br>(0.537-0.801) | 0.292<br>(0.184-0.949) | 0.973<br>(0.822-0.986) | 2.728<br>(1.703-4.370) | 0.184<br>(0.029-1.162) |
| NLR-MuLBSTA | 0.851<br>(0.740-0.963) | 0.837<br>(0.741-0.933) | 1.000<br>(0.631-NA) | 0.679<br>(0.536-0.801) | 0.320<br>(0.205-NA) | 1.000<br>(0.885-1.000) | 3.118<br>(2.107-4.613) | 0.000<br>(0.000-NA) |
| CURB-65 | 0.700<br>(0.505-0.896) | 0.744<br>(0.573-0.915) | 0.625<br>(0.245-0.915) | 0.755<br>(0.617-0.862) | 0.278<br>(0.168-0.712) | 0.930<br>(0.722-0.965) | 2.548<br>(1.247-5.208) | 0.497<br>(0.200-1.232) |
| NLR-CURB-65 | 0.889<br>(0.743-1.036) | 0.870<br>(0.762-0.978) | 0.875<br>(0.473-0.997) | 0.868<br>(0.747-0.945) | 0.500<br>(0.310-0.978) | 0.979<br>(0.855-0.992) | 6.63<br>(3.17-13.86) | 0.144<br>(0.023-0.904) |

AUC, area under curve; SEN, sensitivity; SPE, specificity; PPV, positive predictive value; NPV, negative predictive value; DLR, diagnostic likelihood ratios.

After NLR was incorporated into MuLBSTA (NLR-MuLBSTA) and CURB-65 (NLR-CURB-65) models respectively, it was found that the prediction effect of the improved model was significantly better than that of the original model, but there was no significant difference of AUC in NLR vs. NLR-MuLBSTA and NLR vs. NLR-CURB-65 (p = 0.9675 and p = 0.2971, respectively) (Table 2).

### Performance of the NLR in stratifying risk of patients

The median follow-up time was 10 days, minimum 2 days, and maximum 26 days. Patients were divided into two strata according to the cutoff value of NLR (low risk: < 3.13; high risk: ≥ 3.13) and age (age < 50 years; age ≥ 50 years). Kaplan–Meier analysis showed significant statistical differences in the two groups (p = 0.0005 and p = 0.00028) (Figure 3A and 3B). Furthermore, patients with 2019-nCoV pneumonia were stratified for severe illness incidence with respect to age with NLR (strata 1: age < 50 years and NLR < 3.13; strata 2: age < 50 years and NLR ≥ 3.13; strata 3: age ≥ 50 years and NLR < 3.13; strata 4: age ≥ 50 years and NLR ≥ 3.13). There was no severe illness case in the strata 1 and strata 2, 1 (9.1%) case in the strata 3 and 7 (50%) cases in the strata 4. As shown in Figure 3C, the severe illness incidence was significantly different between strata 3 and 4 (p = 0.0195), but the difference between the strata 2 and 3 was not significant (p = 0.4142).

**Figure 3.**
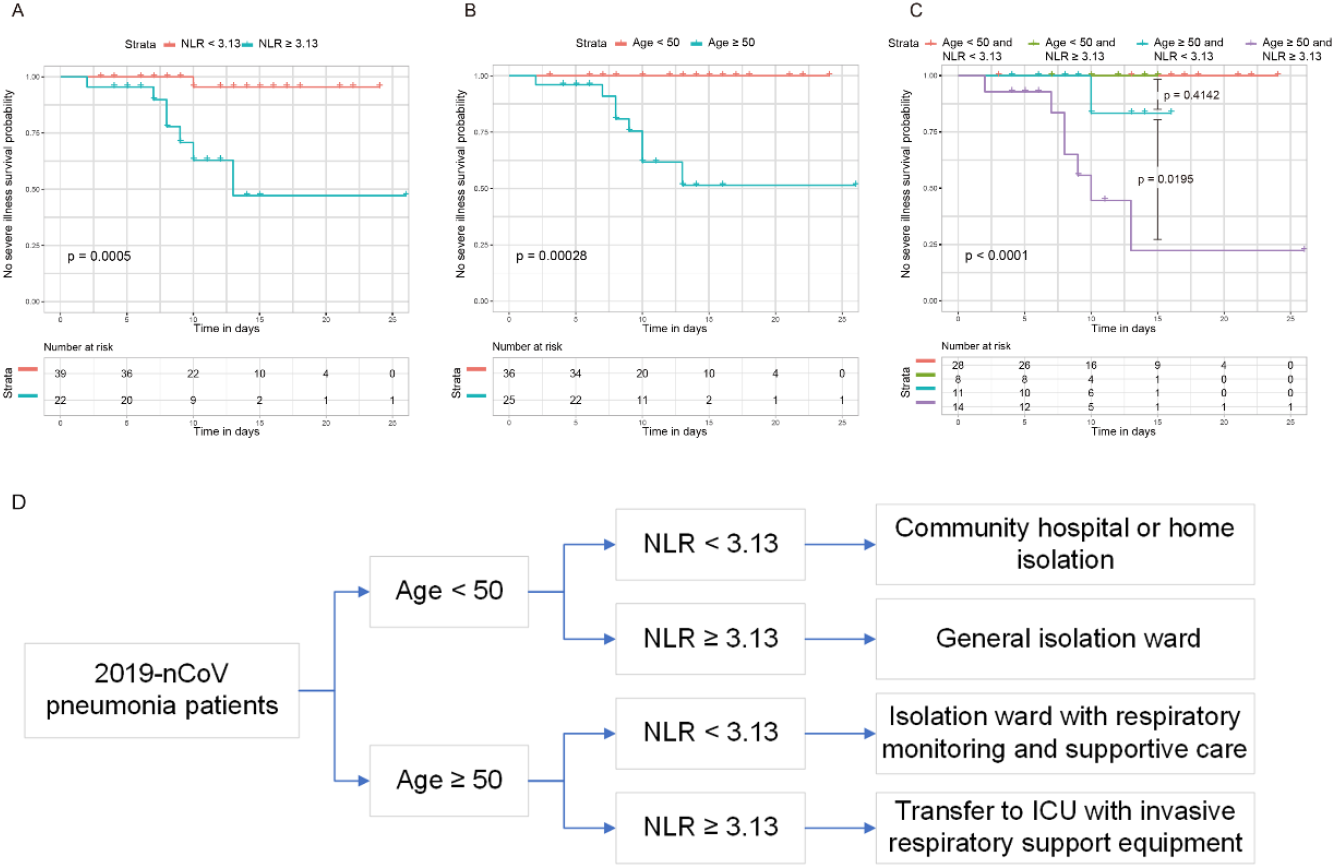
Kaplan–Meier curves of risk group stratification for no severe illness in 2019-nCoV cohorts. (A) The range of NLR for each risk group were as follows: low risk: <3.13, and high risk: ≥ 3.13. (B) Risk group stratification with age and (C) NLR with age. (D) 2019-nCoV pneumonia management process.

## Discussion

Since the outbreak of 2019-nCoV pneumonia in December 2019, there were 2000 to 4000 new confirmed patients every day in China, and the number of severe cases and deaths was also increasing day by day. The research showed that 26% of patients received ICU care, and mortality was 4.3%.^6^ The number of patients in Wuhan and other regions is increasing rapidly. The current difficulty is the shortage of medical resources, especially critical care resources. If severe cases can be identified as early as possible, risk stratification management will help alleviate insufficient medical resources and might reduce mortality. No risk predictive model of severe illness about 2019-nCoV pneumonia has been published to date.

Most of the 2019-nCoV pneumonia patients in the hospital were from Wuhan. Patients with 2019-nCoV pneumonia on early onset were not very severe. The median time from onset to admission of severe patients and mild patients was similar, but the severe patients deteriorated on 7-14 days of illness course and developed to severe pneumonia and acute respiratory failure. The severe or death patients with 2019-nCoV infection were mostly old age with comorbidities.^7^ In the study, severe ill ones are all over 50 years old. The decrease of lymphocyte count was related to the progress of the disease. we proposed that blood lymphocyte related index may be a potential predictor. The NLR was a widely used marker for the assessment of the severity of bacterial infections and the prognosis of patients with pneumonia and tumors. The increase of NLR indicated poor clinical prognosis.^8-14^ The results indicated that 2019-nCoV may act on T lymphocytes, and T lymphocyte damage is an important factor that causes patients to deteriorate^13^. In addition, patients with severe illness might have bacterial infection due to low immune function. The latest report followed up the laboratory test results of patients with 2019-nCoV infection, and found that 5 patients had a rising neutrophil count and a falling lymphocyte count before their deaths.^6^

In the study, the data of 61 patients with 2019-nCoV pneumonia were analyzed, the baseline characteristics of patients in the mild and severe groups were described and compared, and the dynamic changes of laboratory indexes and imaging were demonstrated. The independent risk factors affecting incidence of severe illness were screened. The results showed that NLR was the most significant factor affecting the severe illness incidence, and it had significant predictive value. Furthermore, according to the NLR and age stratification, the incidence of severe ill ones with NLR ≥ 3.13 and aged ≥ 50 years old was 50%, and 9.1% in age ≥ 50 and NLR < 3.13 patients.

Previous studies showed that the MuLBSTA score can early warn the mortality of viral pneumonia, which included six indicators, such as age, smoking history, hypertension, bacterial co-infection, lymphopenia, and multilobular infiltration.^15^ CURB-65 score was widely used to evaluate 30-day mortality of patients with community-acquired pneumonia.^16-18^ In the study, NLR was compared with MuLBSTA and CURB-65 scoring models, the results showed that NLR had higher AUC, c-index, sensitivity and specificity, which indicated that NLR was better than the other two models in early predicting the incidence of severe illness. Furthermore, it was found that the prediction effect of the NLR-MuLBSTA and NLR-CURB-65 models was better than that of the original model. But NLR was an easy-to-use predictor index.

Based on the results, we suggest that patients with 2019-nCoV pneumonia can improve risk stratification and management according to NLR and age model. Patients with age < 50 years old and NLR < 3.13 who are no risk should be treated in a community hospital or home isolation. Patients with age < 50 years old and NLR ≥ 3.13 who are low risk need to be general isolation ward. Patients with age ≥ 50 and NLR < 3.13 patients who are moderate risk, should be admitted to isolation ward with respiratory monitoring and supportive care. Patients with age ≥ 50 and NLR ≥ 3.13 who are high risk should actively transfer to ICU with invasive respiratory support equipment (Figure 3D). If there are large-scale cases, the risk stratification and management will help alleviate the shortage of medical resources and reduce the mortality of critical patients.

There were some limitations in the study. First, the study was a single center with small sample and no external validation cohort. Second, most of patients are still in hospital, whose condition maybe change in follow-up. And the study has not included the final survival outcome. However, we focused on the early identification of critical cases for risk stratification and management. We expect that the risk model can help alleviate the shortage of medical resources and manage the patients with 2019-nCoV pneumonia.

## Conclusion

The NLR was the most useful prognostic factor affecting the prognosis for severe illness patients with 2019-nCoV pneumonia. Early application of NLR with age was beneficial to patients’ classification management and relief of medical resource shortage.

## Data Availability

The data used to support the findings of this study are available from the corresponding author upon request.

## Acknowledgments

We are grateful to Yi Ning, Ph.D, M.D. for his constructive comments in improving the language, grammar, and readability of the paper.

